# Clinical scenario related to cardiovascular system: is it possible to develop thoracic pain imitating a musculoskeletal disorder? A scoping review

**DOI:** 10.1101/2024.02.25.24302970

**Authors:** Nicola De Meo, David Poselek, Michele Margelli, Andrea Segat, Martina Zaninetti, Marco Segat, Federico Minetti, Giovanni Galeoto, Filippo Maselli, Matteo Fascia

**Affiliations:** Department of Human Neuroscience “Sapienza” University of Rome, Roma, Italy; Department of Human anatomy, Histology, Forensic medicine and Orthopedics, “Sapienza” University of Rome, Italy; Department of Morphology Surgery and Experimental Medicine, Ferrara University, Ferrara, Italy; Department of Neuroscience, University of Padova, Padova, Italy; Department of Medicine and Health Science “Vincenzo Tiberio”, University of Molise, Campobasso, Italy; Department of Human Neurosciences, University of Roma “Sapienza Roma”, Rome, Italy; Department of Information Engineering - University of Brescia, Italy

## Abstract

**Background:** When assessing a patient presenting with thoracic pain it’s important for the physiotherapist to quickly understand if the cause is a musculoskeletal condition or a dysfunction of the cardiovascular and circulatory system. Promptly referring the patient is essential to identify potentially life-threatening conditions at an early stage.

**Objectives:** Identifying the current state of knowledge regarding cardiovascular and circulatory systems conditions that generate a thoracic pain that resembles a musculoskeletal condition.

**Study Design:** Scoping review

**Eligibility criteria:** This review will incorporate studies encompassing any research design. Inclusion is limited to articles written in either English or Italian language. Our population of interest specifically includes patients experiencing thoracic pain, with no restrictions regarding age and gender, to ensure a comprehensive understanding of the condition’s impact across different demographics. The concept under investigation is the manifestation of symptoms in the thoracic region, which are attributed to cardiovascular disorders or dysfunctions. It is critical to our scope that we delineate the context by intentionally omitting studies set in emergency contexts. This exclusion criterion allows the review to narrow its focus on the tools employed in making differential diagnoses without relying on instrumental examinations, thus aiming to elucidate diagnostic strategies applicable in a non-emergent setting.

**Results:** 

**Conclusions:** 

## Introduction

### Rationale

Thoracic pain is a common complaint and distinguishing between cardiovascular and musculoskeletal causes is crucial for appropriate treatment and management especially because misdiagnosis or delayed diagnosis of a cardiovascular disorder can lead to serious consequences ^[1-2-3-4]^. Symptoms of cardiovascular and musculoskeletal disorders in the thoracic region can overlap making it challenging for clinicians to differentiate between the two especially for those that operate outside a hospital setting since they may not have access to some specific instrumental examinations.^[3-4]^. A scoping review on this topic could identify gaps in the literature highlight areas that require further research and help the development of guidelines for the differential diagnosis outside the hospital setting.

## Objectives

### Research question

identifying and describing the current state of knowledge regarding the differential diagnosis between musculoskeletal and cardiovascular conditions in patients with thoracic pain. Secondary objectives: Defining all the signs and symptoms that should be accurately investigated; Identifying the tools that support the physiotherapist in the differential diagnosis between a musculoskeletal and cardiovascular thoracic pain.

According to the Joanna Briggs Institute (JBI) guidelines ^[5]^ A “PCC” strategy was used to state the research question (**Population:** Patients with thoracic pain. No limits of age and gender were established; **Concept:** The symptoms in the thoracic region are caused by cardiovascular disorders or dysfunctions; **Context:** We decided to exclude emergency contexts from this review to focus on the tools to make differential diagnosis without instrumental examinations

## Methods

### Protocol and registration

Registration on Medxriv (https://www.medrxiv.org)

### Eligibility criteria

#### Inclusion criteria

any type of study design. No geographical or temporal limits of publication will be applied. Only articles in English and Italian or at least with an English abstract will be considered. All the studies that do not match the PCC described above will be excluded. The rationale of the choice is contained in our initial question.

### Information sources

The research will be conducted on the following databases: Medline, Embase, CINAHL, TRIP Database; we will search for research protocols on PROSPERO and clinicaltrial.gov. We will search for grey literature on opengrey.eu and Google Scholar.

### Search Strategy

As recommended in all JBI types of reviews and PRISMA-S a three-step search strategy will be utilized ^[5]^: The first step will be an initial search of an appropriate online database relevant to the topic (PubMed). This initial search will be followed by an analysis of the text words contained in the title and abstract of retrieved papers and of the index terms used to describe the articles. A background search using all identified keywords and index terms will be undertaken on Pubmed. The reference list of identified reports and articles will be searched for additional sources. No search limitations and filters will be applied except for the language (English and Italian).

Different search terms and keywords will be used, such as “cardiovascular”, “cardiac”, “vascular”, “disease”, “pathology”, “condition”, “disorder”, “thora*”, “chest”, “dorsal”, “back”, “musculoskeletal”, “muscular”, “skelet*”, “articular”, “differential”, “diagnosis”, “screening”, “referral”, “flag*”.

These words will be combined differently according to database functioning.

### Selection of sources of evidence

Selection process is based on title and abstract by two independent reviewers. Disagreements on study selection and data extraction will be discussed with another reviewer if needed. Selection is performed based on inclusion criteria pre-specified above. The software used for the management of the results will be Zotero ^[6]^. For excluded studies reasons should be stated on why they were excluded. There should be a narrative description of the process accompanied by a flowchart of review process (from the PRISMA-ScR statement^[7]^). Pilot testing of source selectors: Random sample of 25 titles/abstracts is selected. The entire team screens these using the eligibility criteria and definitions/elaboration document then team meets to discuss discrepancies and make modifications to the eligibility criteria and definitions/elaboration document. Team only starts screening when 75% (or greater) agreement is achieved. When screening started we’ll use a Microsoft Excel file to manage and catalogue the articles resulting from the search.

### Data charting process

Team trial the extraction from one two or three sources to ensure all relevant results are extracted by at least two members of the review team. A template data extraction instrument for source details characteristics and results extraction is provided in appendix. Pilot step: the extraction form will be tested on two or three sources to ensure all relevant results are extracted by two blinding members of the review team. Inconsistencies were resolved by a third reviewer. This form will be reviewed by the research team and pre-tested by all reviewers before implementation to ensure that the form captures the information accurately modifications will be detailed in the full scoping review.

### Data items

Key informations will described in a charting table with the description of:

- Authors
- Year of publication
- Country of publication
- Study design
- Age
- Predictive factors of a cardiovascular condition
- Signs
- Symptoms
- Localization of the symptoms
- Intensity of the symptoms
- Useful tests for differential diagnosis
- Referrals to other professionists.

### Critical appraisal of individual sources of evidence

No critical appraisal will be performed according to JBI guidelines for Scoping Review^[8]^

### Synthesis of results

The results will be presented as a map of the data extracted from the included papers in a diagrammatic, tabular form, and in a descriptive format that aligns with the objectives and scope of the review. Descriptive analysis: distribution of sources of evidence by year or period of publication, countries of origin, area of intervention (clinical, policy, educational, etc.), and research methods. The results can also be classified under the main conceptual categories such as:

- Features of the signs and symptoms;
- Level of concern;
- Systems involved (musculoskeletal, cardiovascular, gastroenterological, respiratory, psychological);
- Setting

## Data Availability

All data produced in the present work are contained in the manuscript

## Bibliography

1. -ller C, Christ M. The Interdisciplinary Management of Acute Chest Pain. Dtsch Arztebl Int. 2015;112(45):768– 780. doi:10.3238/arztebl.2015.0768;

2. sner S, Haasenri□er J, Becker A, et al. Ruling out coronary artery disease in primary care: development and validation of a simple prediction rule. CMAJ. 2010;182(12):1295– 1300. doi:10.1503/cmaj.100212;

3. Cayley WE. Diagnosing the Cause of Chest Pain. 2005;72(10);

4. Craske MG, Stein MB, Eley TC, et al. Anxiety disorders. Nat Rev Dis Primers. 2017;3:17024. doi:10.1038/nrdp.2017.24;

5. Recommendations for the extraction, analysis, and presentation of results in scoping reviews. Pollock, Danielle ; Peters, Micah D J ; Khalil, Hanan ; McInerney, Patricia ; Alexander, Lyndsay ; Tricco, Andrea C ; Evans, Catrin ; de Moraes, Érica Brandão ; Godfrey, Christina M ; Pieper, Dawid ; Saran, Ashrita ; Stern, Cindy ; Munn, Zachary. JBI evidence synthesis, 2023-03, Vol.21 (3), p.520–532;

6. https://www.zotero.org/;

7. Tricco AC, Lillie E, Zarin W, et al. PRISMA Extension for Scoping Reviews (PRISMA-ScR): Checklist and Explanation. Ann Intern Med. 2018;169(7):467–473. doi:10.7326/M18-0850;

8. Peters MDJ, Godfrey C, McInerney P, Munn Z, Tricco AC, Khalil, H. Chapter 11: Scoping Reviews (2020 version). Aromataris E, Munn Z, editors. JBI Manual for Evidence Synthesis. JBI; 2020.

